# Changes in the behavioural determinants of health during the coronavirus (COVID-19) pandemic: gender, socioeconomic and ethnic inequalities in 5 British cohort studies

**DOI:** 10.1101/2020.07.29.20164244

**Authors:** David Bann, Aase Villadsen, Jane Maddock, Alun Hughes, George B. Ploubidis, Richard J. Silverwood, Praveetha Patalay

## Abstract

**Background:** The coronavirus (COVID-19) pandemic and physical distancing measures are expected to have far-reaching consequences on population health, particularly in already disadvantaged groups. These consequences include changes in health impacting behaviours (such as exercise, sleep, diet and alcohol use) which are important drivers of health inequalities. We sought to add to the rapidly developing empirical evidence base investigating the impacts of the pandemic on such behavioural outcomes.

**Methods:** Using data from five nationally representative British cohort studies (born 2000-2, 1989-90, 1970, 1958, and 1946), we investigated sleep, physical activity (exercise), diet, and alcohol intake (N=14,297). Using measures of each behaviour reported before and during lockdown, we investigated change in each behaviour, and whether such changes differed by age/cohort, gender, ethnicity, and socioeconomic position (SEP; childhood social class, education attainment, and adult reporting of financial difficulties). Binary logistic regression models were used, accounting for study design and incorporating non-response weights, to estimate absolute differences in each outcome before and during lockdown within each cohort. Meta-analysis was used to pool cohort-specific estimates and formally test for heterogeneity across cohorts.

**Results:** Changes in these outcomes occurred in both directions, i.e. shifts from the middle part of the distribution to both declines and increases in sleep, exercise, and alcohol use. For all outcomes, older cohorts were less likely to report changes in behaviours compared with younger cohorts. In the youngest cohort (born 2001), the following shifts were more evident: increases in exercise, fruit and vegetable intake, sleep duration, and less frequent alcohol consumption. Widening inequalities in sleep during lockdown were more frequent amongst females, socioeconomically disadvantaged groups, and ethnic minorities. For other outcomes, inequalities were largely similar before and during lockdown, yet ethnic minorities were increasingly likely during lockdown to undertake less exercise and consume lower amounts of fruit and vegetables.

**Conclusions:** Our findings highlight the multiple changes to behavioural outcomes that may have occurred due to COVID-19 lockdown, and the differential impacts across generation, gender, socioeconomic circumstances across life, and ethnicity. Such changes require further monitoring given their possible implications to population health and the widening of health inequalities.

## Introduction

The coronavirus (COVID-19) pandemic is expected to have far-reaching consequences on population health, particularly in already disadvantaged groups.^1 2^ Aside from direct effects of COVID-19 infection, detrimental changes may include effects on physical and mental health due to associated changes to health-impacting behaviours. Change in such behaviours may be anticipated due to the effects of social distancing, both mandatory and voluntary, and change in factors which may affect such behaviours—such as employment, financial circumstances, and mental distress.^3 4^ The behaviours investigated here include physical activity, diet, alcohol, and sleep^5^—likely key contributors to existing health inequalities^6^ and indirectly implicated in inequalities arising due to COVID-19 given their link with outcomes such as obesity and diabetes.^7^

While empirical evidence of the impact of COVID-19 on such behaviours is emerging,^8-26^ it is currently difficult to interpret for multiple reasons. First, generalising from one study location and/or period of data collection to another is complicated by the vastly different societal responses to COVID-19 which could plausibly impact on such behaviours, such as restrictions to movement, access to restaurants/pubs, and access to support services to reduce substance use. This is compounded by many studies investigating only one health behaviour in isolation. Further, assessment of change in any given outcome is notoriously methodologically challenging.^27^ Some studies have questionnaire instruments which appear to focus only on the negative consequences of COVID-19,^8^ thus curtailing an assessment of both the possible positive and negative effects on health behaviours.

The consequences of COVID-19 lockdown on behavioural outcomes may differ by factors such as age, gender, socioeconomic position (SEP), and ethnicity—thus potentially widening already existing health inequalities. For instance, younger generations (e.g. age 18-30 years) are particularly affected by cessation or disruption of education, loss of employment and income;^3^ and were already less likely than older persons to be in secure housing, secure employment, or stable partnerships.^28^ In contrast, older generations appear more susceptible to severe consequences of COVID-19 infection, and in many countries were recommended to ‘shield’ to prevent such infection. Within each generation, the pandemic’s effects may have had inequitable effects by gender (e.g. childcare responsibilities being borne more by women), SEP and ethnicity (e.g. more likely to be in at-risk and low paid employment, insecure and crowded housing).

Using data from five nationally representative British cohort studies, which each used an identical COVID-19 follow-up questionnaire in May 2020, we investigated change in multiple health-impacting behaviours. Multiple outcomes were investigated since each is likely to have independent impacts on population health, and evidence-based policy decisions are likely better informed by simultaneous consideration of multiple outcomes.^29^ We considered multiple well-established health equity stratifiers:^30^ age/cohort, gender, socioeconomic position (SEP) and ethnicity. Further, since childhood SEP may impact on adult behaviours and health outcomes independently of adult SEP,^31^ we utilised previously collected prospective data in these cohorts to investigate childhood and adult SEP.

## Methods

### Study samples

We used data from four British birth cohort (c) studies, born in 1946,^32^ 1958,^33^ 1970,^34^ and 2000-2002 (born 2000-2; 2001c, inclusive of Northern Ireland);^35^ and one English longitudinal cohort study (born 1989-90; 1990c) followed up from 14 years.^36^ Each has been followed up at regular intervals from birth or adolescence; on heath, behavioural, and socioeconomic factors. In each study, participants gave written consent to be interviewed. Research ethics approval was obtained from relevant committees. In May 2020, during the COVID-19 pandemic, participants were invited to take part in an online questionnaire which measured demographic factors, health measures and multiple behaviours.^37^

### Outcomes

We investigated the following behaviours: sleep (number of hours each night on average), exercise (number of days per week (i.e. from 0-7) the participants exercised for 30 mins or more at moderate-vigorous intensity—“working hard enough to raise your heart rate and break into a sweat”), and diet (number of portions of fruit & vegetables per day (from 0 to ≥6; portion guidance was provided). Alcohol consumption was reported in both consumption frequency (never to 4 or more times per week) and the typical number of drinks consumed when drinking (number of drinks per day); these were combined to form a total monthly consumption. For each behaviour, participants retrospectively reported levels in “the month before the coronavirus outbreak” and then during the fieldwork period (May 2020). Herein, we refer to these reference periods as pre and during lockdown, respectively. In subsequent regression modelling, binary outcomes were created for all outcomes, chosen to capture high-risk groups in which there was sufficient variation across all cohort and risk factor subgroups—sleep (1=<6 hours or >9 hours per night given its non-linear relation with health outcomes^38 39^), exercise (1=2 or fewer days/week exercise), diet (1=2 or fewer potions of fruit and vegetables/day), alcohol (1=≥14 drinks per week or 5 or more drinks per day; 0=lower frequency and/or consumption).^40^

### Risk factors

Socioeconomic position was indicated by childhood social class (at 10-14 years old), using the Registrar General’s Social Class scale— I (professional), II (managerial and technical), IIIN (skilled non-manual), IIIM (skilled manual), IV (partly-skilled), and V (unskilled) occupations. Highest educational attainment was also used, categorised into four groups as follows: degree/higher, A levels/diploma, O Levels/GCSEs, or none (for 2001c we used parents’ highest education as many were still undertaking education). Financial difficulties were based on whether individuals (or their parents for 2001c) reported (prior to COVID-19) as managing financially comfortably, all right, just about getting by, and difficult. These ordinal indicators were converted into cohort-specific ridit scores to aid interpretation—resulting in relative or slope indices of inequality when used in regression models (ie, comparisons of the health difference comparing lowest with highest SEP).^41^ Ethnicity was recorded as White and non-White—with analyses limited to the 1990c and 2001c owing to a lack of ethnic diversity in older cohorts.

### Statistical analyses

We calculated average levels and distributions of each outcome pre and during lockdown. Logistic regression models were used to examine how gender, ethnicity and SEP were related to each outcome, both before and during lockdown. Where the prevalence of the outcome differs across time, comparing results on the relative scale can impair comparisons of risk factor-outcome associations (eg, identical odds ratios can reflect different associations on the absolute scale).^42^ Thus, we estimated absolute (risk) differences in outcomes by gender, SEP and ethnicity (the *margins* command in Stata following logistic regression). Models examining ethnicity and SEP were gender-adjusted. We conducted cohort-specific analyses and conducted meta-analyses to assess pooled associations, formally testing for heterogeneity across cohorts (I^2^ statistic). To understand the changes which led to differing inequalities we also tabulated calculated change in each outcome (decline, no change, and increase) by each cohort and risk factor group. To confirm that the patterns of inequalities observed using binary outcomes was consistent with results using the entire distribution of each outcome, we additionally tabulated all outcome categories by cohort and risk factor group.

To account for possible bias due to missing data, we weighted our analysis using weights constructed from logistic regression models—the outcome was response during the COVID-19 survey, and predictors were demographic, socioeconomic, household, and individual-based predictors of non-response at earlier sweeps, based on previous work in these cohorts.^43 44 37^ We also used weights to account for the stratified survey designs of the 1946c, 1990c, and 2001c. Stata v15 (STATA corp) was used to conduct all analyses.

## Results

Cohort-specific responses were as follows: 1946c: 1258 of 1843 (68%); 1958c: 5178 of 8943 (58%), 1970c: 4223 of 10458 (40%); 1990c: 1907 of 9380 (20%); 2001c: 2645 of 9946 (27%). The following factors, measured in prior data collections, were associated with increased likelihood of response in this COVID-19 dataset: being female, higher education attainment, higher household income, and more favourable self-rated health. Valid outcome data were available in both pre and during lockdown periods for the following: sleep, N=14,171; exercise, N=13,997; alcohol, N=14,297; fruit/vegetables, N=13,623.

### Overall changes and cohort differences

Outcomes pre and during lockdown were each moderately-highly positively correlated—Spearman’s R as follows: sleep=0.55, exercise=0.58, alcohol=0.76, fruit/vegetable consumption=0.81. For all outcomes, older cohorts were less likely to report change in behaviour compared with younger cohorts (Supplementary Table 1).

The average (mean) amount of sleep (hours per night) was either similar or slightly higher during compared with before lockdown. In each cohort, the variance was higher during lockdown (Table 1)—this reflected the fact that more participants reported either reduced or increased amounts of sleep during lockdown (Figure 1). In 2001c compared with older cohorts, more participants reported increased amounts of sleep during lockdown (Figure 1 and Supplementary Tables 1-2). Mean exercise frequency levels were similar during and before lockdown (Table 1). As with sleep levels, the variance was higher during lockdown, reflecting both reduced and increased amounts of exercise during lockdown (Figure 1 and Supplementary Table 2). In 2001c, a larger fraction of participants reported transitions to no alcohol consumption during lockdown than in older cohorts (Supplementary Table 2). Fruit and vegetable intake was broadly similar pre and during lockdown, although increases in consumption were most frequent in 2001c compared with older cohorts (Figure 1 and Supplementary Table 1).

**Figure 1.**
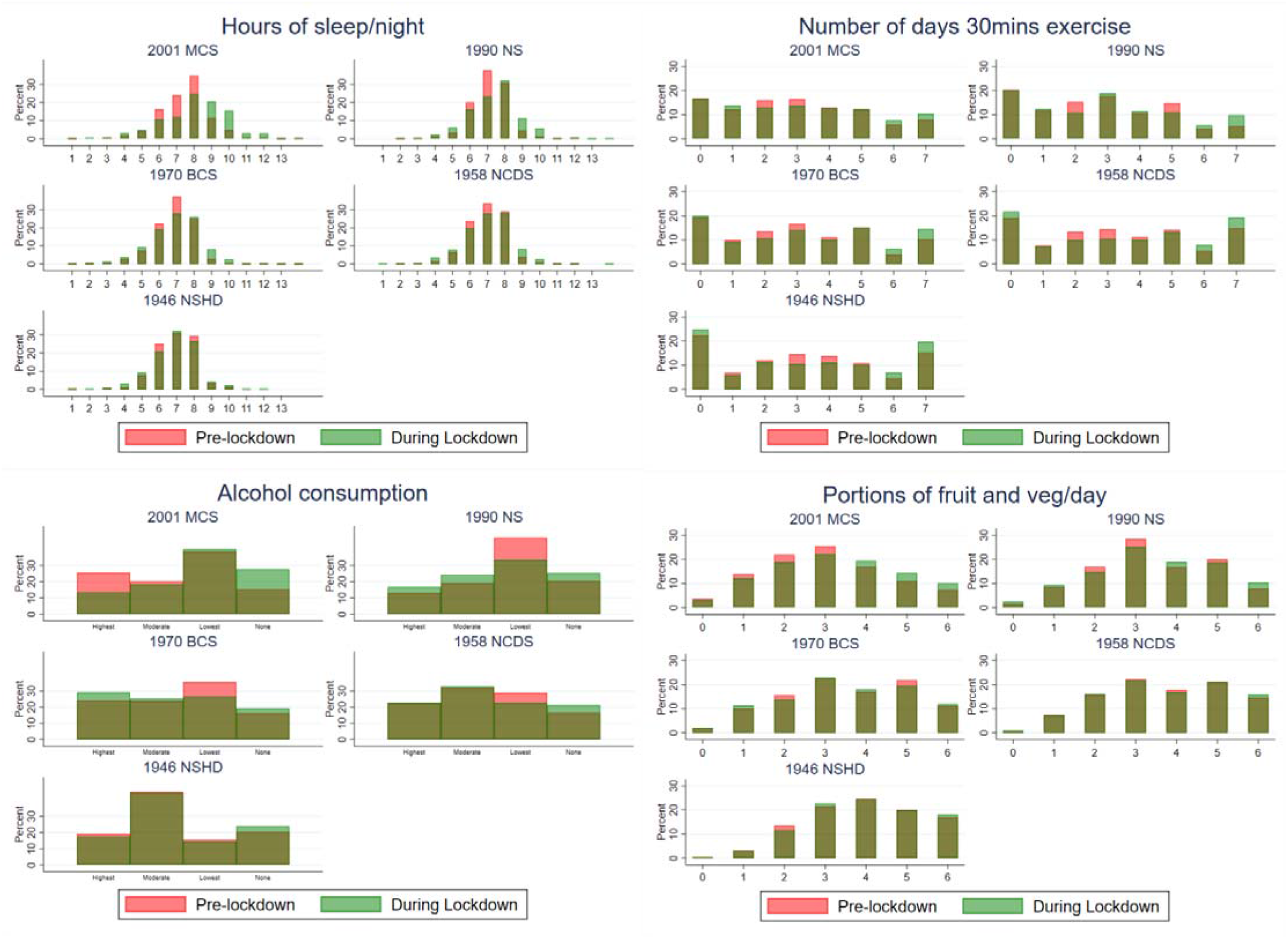
Pre- and during COVID-19 lockdown distributions of health-related behaviours, by cohort. Note: dark green shows overlap, estimates are weighted to account for survey non-response; alcohol consumption was derived as >36, 16-36, 1-15, no drinks per month.

**Table 1.** Participant characteristics: data from 5 British cohort studies.

|  | Cohort study, birth year |  |  |  |  |
| --- | --- | --- | --- | --- | --- |
|  | 2001 | 1990 | 1970 | 1958 | 1946 |
| **Cohort characteristics and risk factors** |  |  |  |  |  |
| Sample size, N | 2164 | 1661 | 3804 | 4574 | 1080 |
| Age in years | 19-20 | 30-31 | 50 | 62 | 74 |
| Males (%) | 49.4% | 43.6% | 51.3% | 50.9% | 50.4% |
| Father's social class, % manual | 23.1% | 37.7% | 58.3% | 63.1% | 67.3% |
| Education attainment, % GCSEs-none | 47.3% | 35.9% | 46.6% | 49.2% | 70.8% |
| Financial difficulties, % difficult | 18.1% | 16.1% | 21.0% | 13.4% | 4.3% |
| **Outcomes** |  |  |  |  |  |
| Pre: Sleep (# hours/day), mean (SD) | 7.5 (1.4) | 7.1 (1.1) | 6.8 (1.2) | 7.0 (1.2) | 6.9 (1.2) |
| During: Sleep (# hours/day), mean (SD) | 8.1 (1.9) | 7.4 (1.5) | 6.9 (1.5) | 7.0 (1.4) | 6.9 (1.3) |
| Pre: Sleep, % atypical (<6 >9 hrs/night) | 12.9% | 6.9% | 12.0% | 10.0% | 10.8% |
| During: Sleep, % atypical (<6 >9 hrs/night) | 31.7% | 16.5% | 18.4% | 15.6% | 16.1% |
| Pre: Exercise (#days/week), mean (SD) | 3.0 (2.1) | 2.7 (2.1) | 3.0 (2.2) | 3.3 (2.4) | 3.2 (2.4) |
| During: Exercise (#days/week), mean (SD) | 3.1 (2.3) | 2.9 (2.2) | 3.3 (2.4) | 3.5 (2.6) | 3.3 (2.6) |
| Pre: Exercise (% 0-2 days/week) | 28.8% | 32.0% | 29.7% | 26.9% | 29.5% |
| During: Exercise (% 0-2 days/week) | 30.4% | 32.5% | 29.2% | 29.1% | 30.8% |
| Pre: Alcohol intake, % never | 15.3% | 20.5% | 16.1% | 16.2% | 18.5% |
| During: Alcohol intake, % never | 27.7% | 25.2% | 19.0% | 20.9% | 20.7% |
| Pre: Alcohol intake, % high risk | 32.6% | 16.0% | 17.3% | 17.2% | 14.2% |
| During: Alcohol intake, % high risk | 13.0% | 12.7% | 21.7% | 17.4% | 14.6% |
| Pre: Fruit/veg intake (# portions), mean (SD) | 3.0 (1.5) | 3.4 (1.5) | 3.5 (1.6) | 3.7 (1.5) | 4.0 (1.4) |
| During: Fruit/veg intake (# portions), mean (SD) | 3.2 (1.6) | 3.5 (1.6) | 3.5 (1.6) | 3.7 (1.6) | 4.0 (1.4) |
| Pre: Fruit/veg intake (% 0-2 portions) | 39.4% | 27.0% | 27.4% | 24.3% | 16.3% |
| During: Fruit/veg intake (% 0-2 portions) | 34.4% | 26.9% | 27.3% | 24.5% | 14.7% |

Note: estimates are weighted to account for survey non-response. High risk drinking is consuming more than 14 drinks a week or more than 5 drinks in a typical drinking day.

### Gender inequalities

Females had a higher risk than males of atypical sleep levels (ie, <6 or >9 hours), and such differences were larger during compared with before lockdown (pooled percent risk difference during (males vs females, during lockdown: −4.2 (−6.6, −2.0), before: −1.9 (−3.7, −0.2); Figure 2). These differences were similar in each cohort (I^2^ =11.6% and 0%, respectively), and reflected greater change in female sleep levels during lockdown (Supplementary Table 1). Before lockdown, in all cohorts females undertook less exercise than males; during lockdown, this difference reverted to null (Figure 2). This was due to relatively more females reporting increased exercise levels during lockdown compared with before (Supplementary Table 1). Males had higher alcohol consumption than females, and reported lower fruit and vegetable intake; effect estimates were slightly weaker during compared with before lockdown (Figure 2).

**Figure 2.**
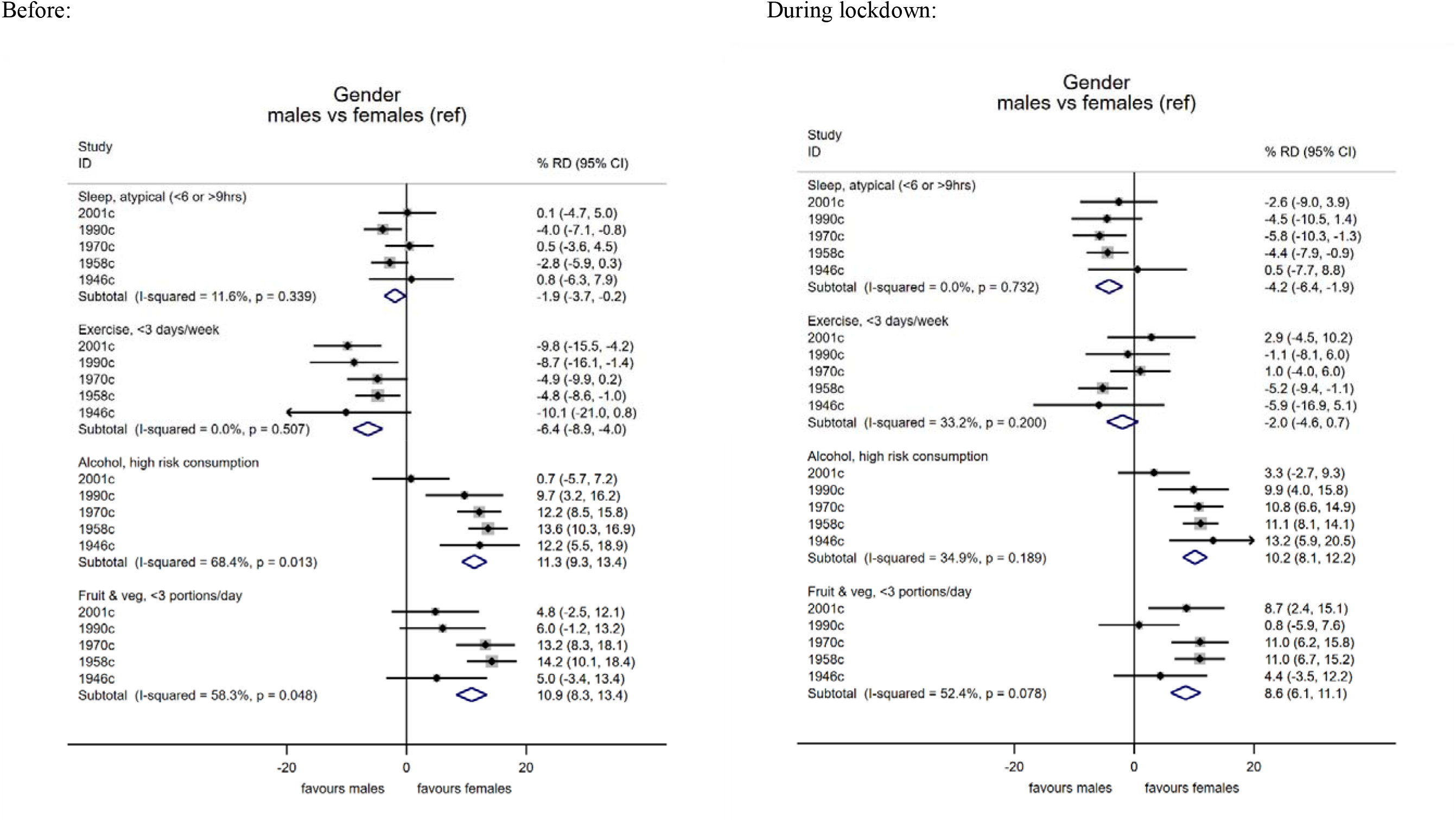
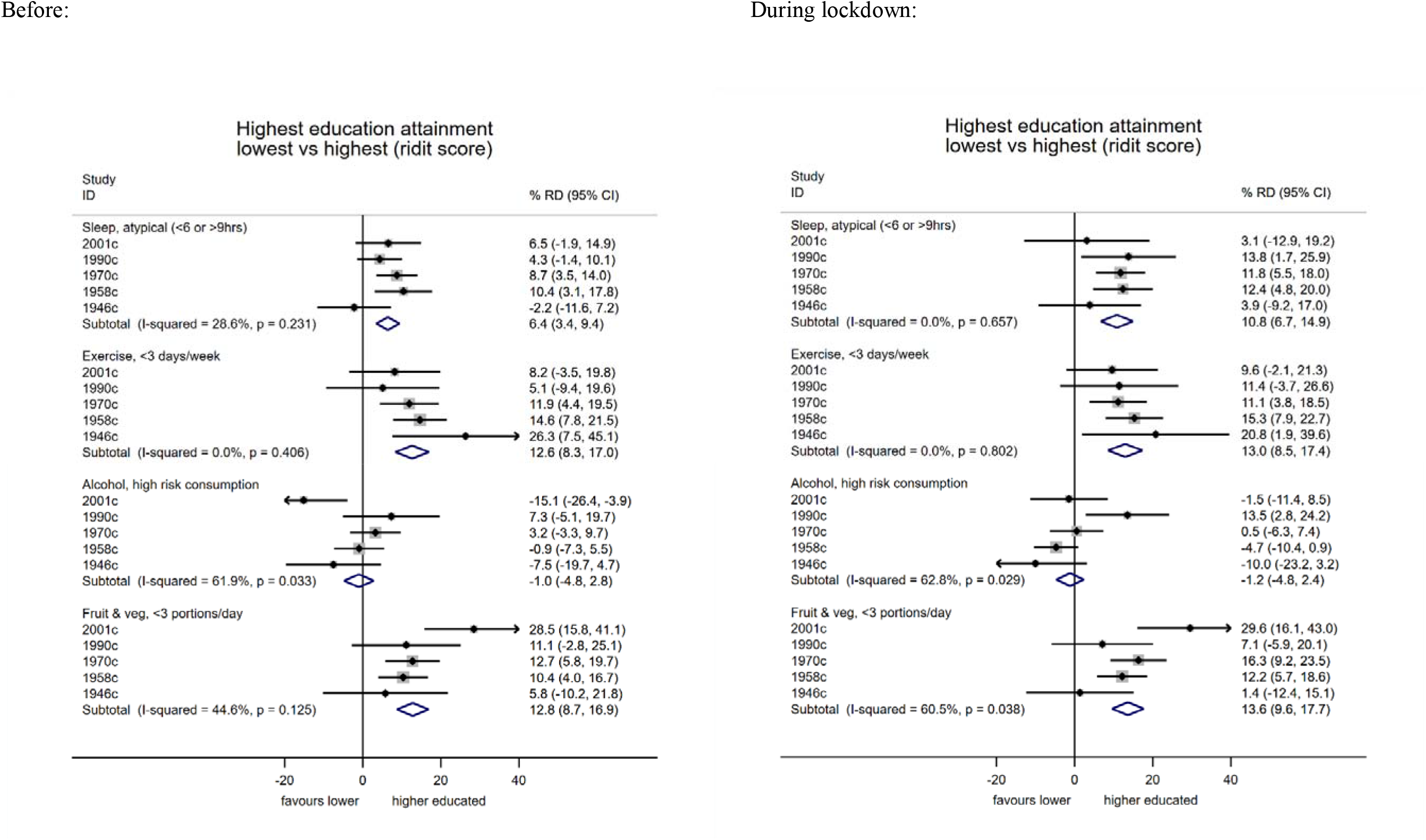
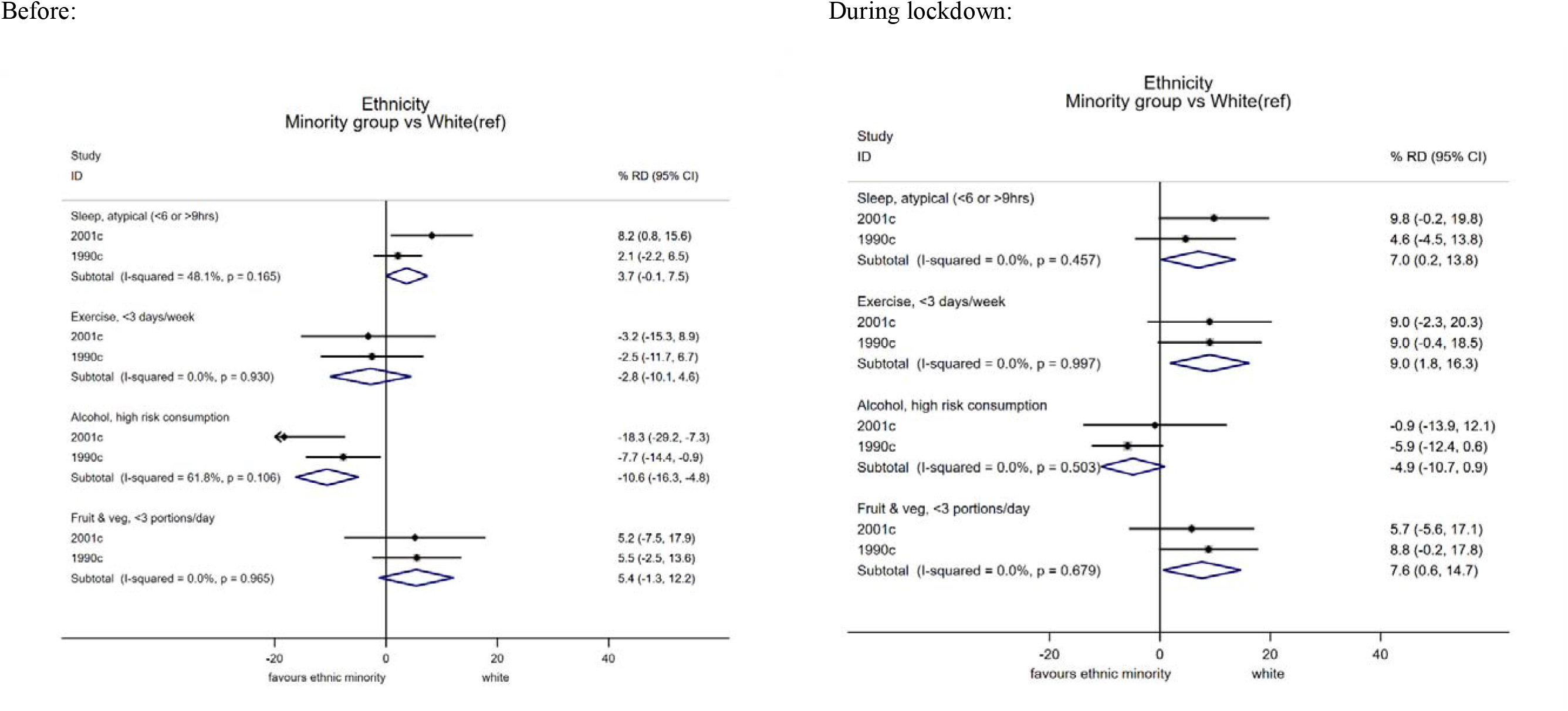
Differences in multiple health behaviours during COVID-19 lockdown (May 2020) compared with prior levels, according to gender, education attainment, and ethnicity: a meta-analysis of 5 cohort studies. Note: estimates show the risk difference on the percentage scale and are weighted to account for survey non-response; ridit scores represent the odds ratio of the least compared with most disadvantaged socioeconomic position).

### Socioeconomic inequalities

Those with lower education had higher risk of atypical sleep levels—this difference was larger and more consistently found across cohorts during compared with before lockdown (Figure 2). Lower education was also associated with lower exercise participation, and with lower fruit and vegetable intake (particularly strongly in 2001c), but not with alcohol consumption; estimates of association were similar before and during lockdown (Figure 2). Associations of childhood social class and adulthood financial difficulties with these outcomes were broadly similar to those for education attainment (Supplementary Figure 1)— differences in sleep during lockdown were larger, and lower childhood social class was more strongly related to lower exercise participation during lockdown (Supplementary Figure 1), and with lower fruit and vegetable intake (particularly in 2001c).

### Ethnic inequalities

Ethnic minorities had higher odds of atypical sleep levels than white participants, with larger effect sizes during compared with before lockdown (Figure 2 and Supplementary Table 1). Ethnic minorities had lower exercise levels during but not before lockdown—pooled percent risk difference during (ethnic minority vs white): 9.0 (1.8, 16.3; I^2^=0%; Figure 2). Ethnic minorities also had higher risk of lower fruit and vegetable intake, with stronger associations during lockdown (Figure 2). In contrast, ethnic minorities had lower alcohol consumption, with stronger effect sizes before lockdown than during (Figure 2).

## Discussion

### Main findings

Using data from 5 national British cohort studies, we estimated the change in multiple health behaviours between pre and during COVID-19 lockdown periods in the UK (May 2020). Where change in these outcomes was identified, it occurred in both directions—ie, shifts from the middle part of the distribution to both declines and increases in sleep, exercise, and alcohol use. In the youngest cohort (2001c), the following shifts were more evident: increases in exercise, fruit and vegetable intake, and sleep, and reduced alcohol consumption frequency. Across all outcomes, older cohorts were less likely to report changes in behaviour. Our findings suggest—for most outcomes measured—a potential widening of inequalities in health-impacting behavioural outcomes which may have been caused by the COVID-19 lockdown.

### Comparison with other studies

In our study the youngest cohort reported increases in sleep during lockdown—similar findings of increased sleep have been reported in many,^13 17 18 24^ but not all^8^ previous studies. Both too much and too little sleep may reflect, and be predictive of, worse mental and physical health.^38 39^ In this sense, the increasing dispersion in sleep we observed may reflect the negative consequences of COVID-19 and lockdown. Females, those of lower SEP, and ethnic minorities were all at higher risk of atypical sleep levels. It is possible that lockdown restrictions and subsequent increases in stress—related to health, job, and family concerns—have affected sleep across multiple generations and potentially exacerbated such inequalities. Indeed, recent work using household panel data in the UK has observed marked increases in anxiety and depression in the UK during lockdown that were largest amongst younger adults.^4^

Our findings on exercise add to an existing but somewhat mixed evidence base. Some studies have reported declines in both self-reported^12 23^ and accelerometery-assessed physical activity,^19^ yet this is in contrast to others which report an increase,^22^ and there is corroborating evidence for increases in some forms of physical activity since online searches for exercise and physical activity appear to have increased.^21^ As in our study, another also reported that males had lower exercise levels during lockdown.^20^ While we cannot be certain that our findings reflect all changes to physical activity levels—lower intensity exercises were not assessed nor was activity in other domains such as in work or travel—the widening inequalities in ethnic minority groups may be a cause of public health concern.

As for the impact of the lockdown on alcohol consumption, concern was initially raised over the observed rises in alcohol sales in stores at the beginning of the pandemic in the UK^45^ and elsewhere. Our findings suggest decreasing consumption particularly in the younger cohort. Existing studies appear largely mixed, some suggesting increases in consumption,^9 16 26^ with others reporting decreases;^11 12 23 25^ others also report increases, yet use instruments which appear to particularly focus on capturing increases and not declines.^8 10^ Different methodological approaches and measures used may account for inconsistent findings across studies, along with differences in the country of origin and characteristics of the sample. The closing of pubs and bars and associated reductions in social drinking likely underlies our finding of declines in consumption amongst the youngest cohort. Increases in fruit and vegetable consumption observed in this cohort may have also reflected the considerable social changes attributable to lockdown, including more regular food consumption at home. However, in our study only positive aspects of diet (fruit and veg consumption) were captured—we did not capture information on volume of food, snacking and consumption of unhealthy foods. Indeed, one study reported simultaneous increases in consumption of fruit and vegetables and snacks.^11^

Further research using additional waves of data collection is required to empirically investigate if the changes and inequalities observed in the current study persist into the future. If the changes persist and/or widen, given the relevance of these behaviours to a range of health outcomes including chronic conditions, COVID-19 infection consequences and years of healthy life lost, the public health implications of these changes may be long-lasting.

### Methodological considerations

While our analyses provide estimates of change in multiple important outcomes, findings should be interpreted in the context of the limitations of this work, with fieldwork necessarily undertaken rapidly. First, self-reported measures were used—while the two reference periods for recall were relatively close in time, comparisons of change in behaviour may have been biased by measurement error and reporting biases. Further, single measures of each behaviour were used which do not fully capture the entire scope of the health-impacting nature of each behaviour. For example, exercise levels do not capture less intensive physical activities, nor sedentary behaviour; while fruit and vegetable intake is only one component of diet. As in other studies investigating changes in such outcomes, we are unable to separate out change attributable to COVID-19 lockdown from other causes—these may include seasonal differences (eg, lower physical activity levels in the pre-COVID-19 winter months), and other unobserved factors which we were unable to account for. If these factors affected the sub-groups we analysed (gender, SEP, ethnicity) equally, our analysis of risk factors of change would not be biased due to this. We acknowledge that quantifying change and examining its determinants is notoriously methodologically challenging—such considerations informed our analytical approach (eg, to avoid spurious associations, we did not adjust for ‘baseline’ (pre-lockdown) measures when examining outcomes during lockdown^46^).

As in other web surveys,^4^ response rates were generally low—while the longitudinal nature of the cohorts enable predictors of missingness to be accounted for (via sample weights),^43 44^ we can’t fully exclude the possibility of unobserved predictors of missing data influencing our results. Finally, we investigated ethnicity using a binary categorisation to ensure sufficient sample sizes for comparisons—we were likely underpowered to investigate differences across the multiple diverse ethnic groups which exist. This warrants future investigation given the substantial heterogeneity within these groups and likely differences in behavioural outcomes.

### Conclusion

Our findings highlight the multiple changes to behavioural outcomes that may have occurred due to COVID-19 lockdown, and the differential impacts—across generation, gender, socioeconomic disadvantage (in early and adult life) and ethnicity. Such changes require further monitoring given their possible implications to population health and the widening of health inequalities.

## Data Availability

2001c, 1990c, 1970c and 1958c data are available from the UK Data Archive: https://www.data-archive.ac.uk. 1946c data are available from: https://www.nshd.mrc.ac.uk/data

https://beta.ukdataservice.ac.uk/datacatalogue/studies/study?id=8658#!/documentation

https://beta.ukdataservice.ac.uk/datacatalogue/studies/study?id=8658#!/details

## Acknowledgements

We thank the Survey, Data, and Administrative teams at the Centre for Longitudinal Studies and Unit for Lifelong Health and Ageing, UCL, for enabling the rapid COVID-19 data collection to take place. We also thanks Professors Rachel Cooper and Mark Hamer for helpful discussions during the COVID-19 questionnaire design period. DB is supported by the Economic and Social Research Council (grant number ES/M001660/1), DB and AV are supported by The Academy of Medical Sciences / Wellcome Trust (“Springboard Health of the Public in 2040” award: HOP001/1025).

## Data availability

**Supplementary Figure 1.**
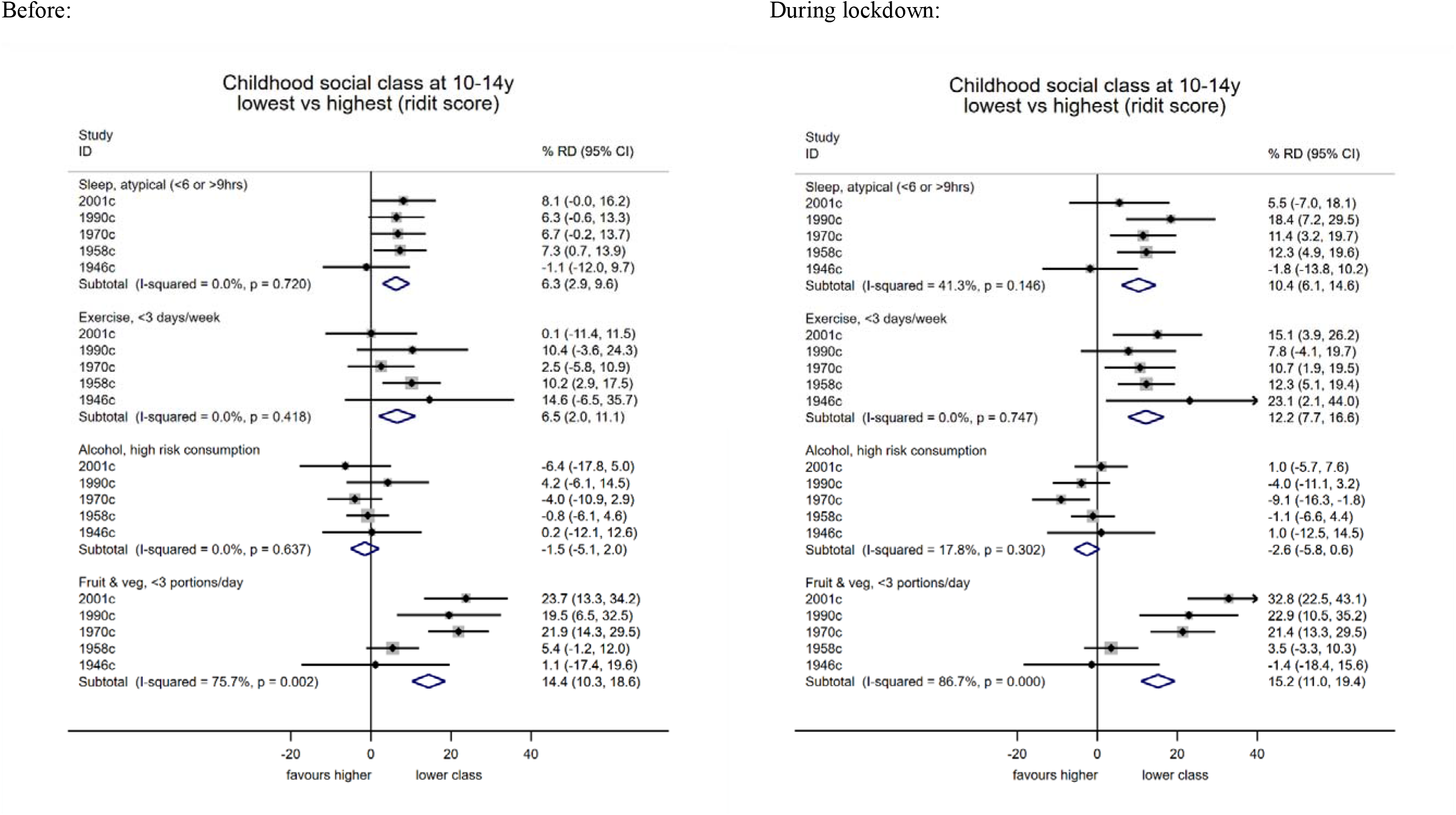
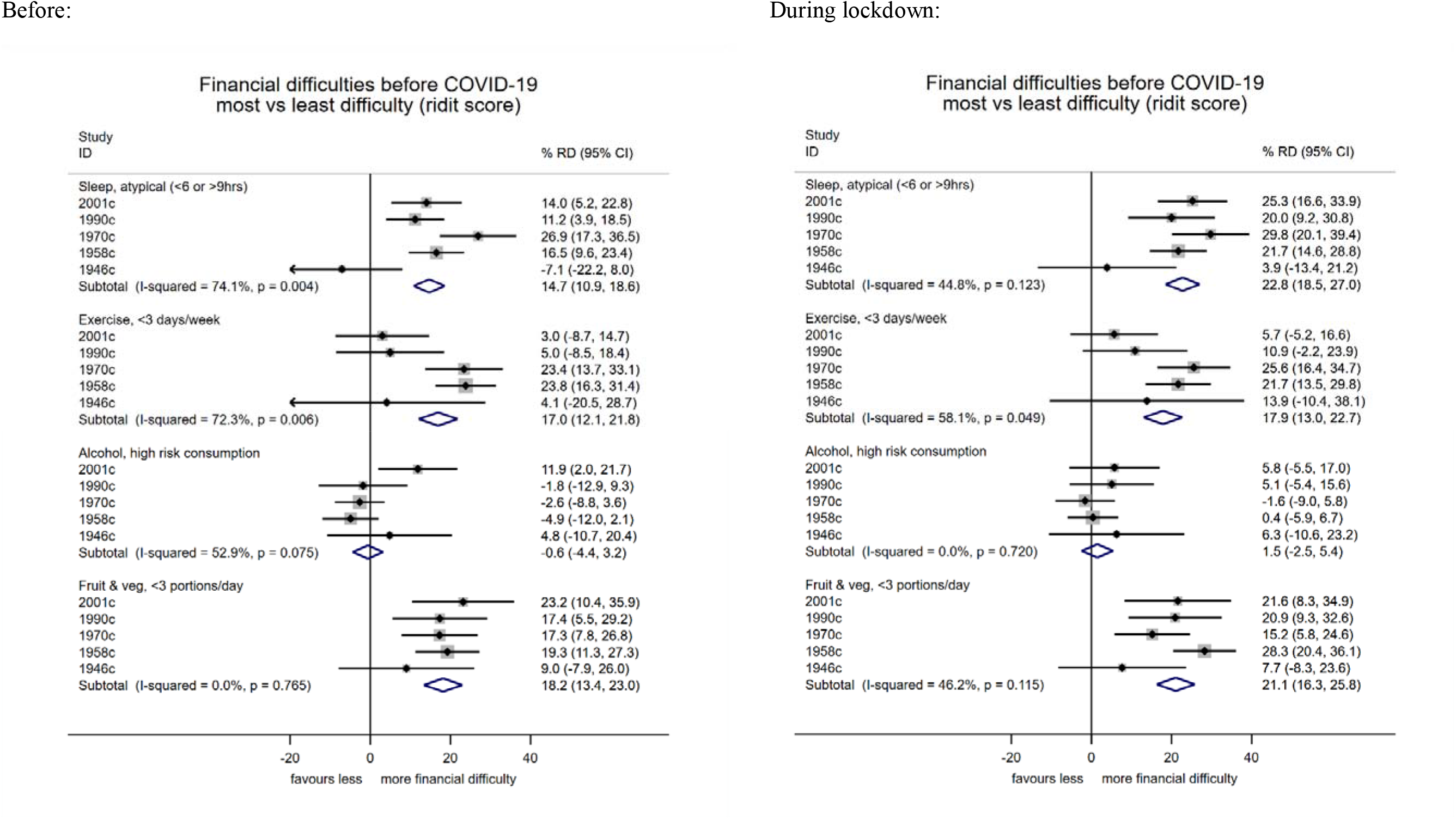
Differences in multiple health behaviours during COVID-19 lockdown (May 2020) compared with prior levels, according to gender, education attainment, and ethnicity: a meta-analysis of 5 cohort studies. Note: estimates show the risk difference on the percentage scale and are weighted to account for survey non-response; ridit scores represent the odds ratio of the least compared with most disadvantaged socioeconomic position, the relative index of inequality)

**Supplementary Table 1.** Changes in behaviors before and during COVID-19 lockdown: percentages in each cohort and risk factor group.

1) Risk factors according to all cohorts:
| **Outcome**, reported change during COVID-19 lockdown | <u>Cohort</u> |  |  |  |  | <u>Gender</u> |  | <u>Education attainment</u> |  | <u>Ethnicity</u> |  |
| --- | --- | --- | --- | --- | --- | --- | --- | --- | --- | --- | --- |
|  | 2001 | 1990 | 1970 | 1958 | 1946 | Females | Males | High education (degree/higher) | Low education (GCSE/none) | White | Non-white |
| **Sleep** |  |  |  |  |  |  |  |  |  |  |  |
| Less sleep | 25.2 | 21.6 | 19.5 | 16.2 | 13.0 | 22.5 | 14.6 | 17.3 | 19.7 | 23.0 | 28.6 |
| no change | 23.0 | 41.3 | 53.9 | 61.2 | 76.5 | 48.0 | 59.3 | 50.2 | 57.6 | 32.0 | 22.7 |
| More | 51.8 | 37.1 | 26.5 | 22.5 | 10.4 | 29.5 | 26.1 | 32.5 | 22.7 | 45.0 | 48.7 |
| **Exercise** |  |  |  |  |  |  |  |  |  |  |  |
| Less exercise | 33.7 | 29.5 | 20.2 | 18.8 | 18.1 | 21.7 | 23.5 | 23.2 | 21.2 | 30.8 | 39.8 |
| no change | 30.0 | 35.3 | 49.1 | 56.2 | 59.8 | 45.7 | 51.2 | 42.7 | 54.3 | 32.2 | 32.2 |
| More | 36.3 | 35.2 | 30.7 | 25.1 | 22.1 | 32.6 | 25.3 | 34.2 | 24.5 | 37.0 | 28.1 |
| **Alcohol** |  |  |  |  |  |  |  |  |  |  |  |
| More alcohol | 16.7 | 30.1 | 24.4 | 15.4 | 10.7 | 21.4 | 16.2 | 20.8 | 17.3 | 23.6 | 13.5 |
| no change | 47.5 | 55.1 | 67.0 | 72.7 | 80.1 | 63.0 | 69.9 | 62.6 | 68.6 | 48.7 | 65.2 |
| Less | 35.8 | 14.8 | 8.6 | 11.9 | 9.2 | 15.6 | 13.9 | 16.6 | 14.1 | 27.6 | 21.3 |
| **Fruit & Veg** |  |  |  |  |  |  |  |  |  |  |  |
| Less fruit & veg | 16.3 | 16.4 | 13.9 | 10.4 | 5.9 | 14.5 | 9.7 | 12.6 | 11.3 | 15.4 | 22.9 |
| no change | 51.7 | 63.0 | 70.0 | 76.6 | 82.3 | 66.8 | 74.2 | 68.7 | 73.4 | 57.7 | 48.7 |
| More | 32.0 | 20.6 | 16.2 | 13.1 | 11.8 | 18.7 | 16.0 | 18.8 | 15.3 | 26.9 | 28.4 |

Note: estimates are weighted to account for survey non-response.

2) Risk factors separately in each cohort
| Cohort, outcome | reported change during COVID-19 lockdown | <u>Gender</u> |  | <u>Education attainment</u> |  | <u>Ethnicity</u> |  |
| --- | --- | --- | --- | --- | --- | --- | --- |
|  |  | Females | Males | High education (degree/higher) | Low education (GCSE/none) | White | Non-white |
| **2001, sleep** | Less sleep | 26.2 | 24.2 | 21.6 | 28.5 | 24.0 | 34.1 |
|  | no change | 20.6 | 25.4 | 22.9 | 23.9 | 24.3 | 14.5 |
|  | More | 53.3 | 50.4 | 55.5 | 47.5 | 51.7 | 51.4 |
| **2001, exercise** | Less exercise | 28.7 | 38.8 | 31.7 | 37.1 | 32.4 | 42.6 |
|  | no change | 28.0 | 32.0 | 29.4 | 29.6 | 30.3 | 27.6 |
|  | More | 43.3 | 29.2 | 38.9 | 33.2 | 37.3 | 29.8 |
| **2001, alcohol** | More alcohol | 17.0 | 16.4 | 16.1 | 18.2 | 17.7 | 9.7 |
|  | no change | 43.5 | 51.7 | 43.2 | 51.4 | 45.4 | 62.9 |
|  | Less | 39.5 | 32.0 | 40.7 | 30.3 | 36.9 | 27.5 |
| **2001, fruit & veg** | Less fruit & veg | 16.3 | 16.4 | 16.2 | 15.7 | 15.0 | 25.2 |
|  | no change | 47.4 | 56.2 | 48.8 | 57.6 | 52.8 | 42.8 |
|  | More | 36.3 | 27.4 | 35.0 | 26.7 | 32.2 | 32.0 |
| **1990, sleep** | Less sleep | 23.2 | 19.7 | 18.8 | 26.4 | 21.7 | 21.2 |
|  | no change | 38.0 | 45.5 | 42.3 | 39.4 | 42.4 | 33.7 |
|  | More | 38.8 | 34.9 | 38.9 | 34.2 | 35.9 | 45.1 |
| **1990, exercise** | Less exercise | 23.7 | 36.7 | 27.3 | 34.4 | 28.5 | 36.0 |
|  | no change | 38.2 | 31.8 | 33.0 | 38.4 | 34.9 | 38.2 |
|  | More | 38.2 | 31.5 | 39.7 | 27.2 | 36.6 | 25.8 |
| **1990, alcohol** | More alcohol | 31.7 | 28.1 | 30.3 | 31.1 | 31.7 | 18.7 |
|  | no change | 54.5 | 56.0 | 54.8 | 54.5 | 53.3 | 68.4 |
|  | Less | 13.9 | 16.0 | 14.9 | 14.4 | 15.1 | 12.9 |
| **1990, fruit & veg** | Less fruit & veg | 18.5 | 14.0 | 16.2 | 13.8 | 15.9 | 20.3 |
|  | no change | 61.7 | 64.6 | 63.6 | 64.4 | 64.0 | 55.6 |
|  | More | 19.8 | 21.5 | 20.2 | 21.9 | 20.1 | 24.1 |
|  |  | Females | Males | High education (degree/higher) | Low education (GCSE/none) | White | Non-white |
| **1970, sleep** | Less sleep | 25.4 | 13.8 | 18.1 | 20.8 | 14.5 | 13.6 |
|  | no change | 46.2 | 61.5 | 51.6 | 57.1 | 69.7 | 72.0 |
|  | More | 28.4 | 24.7 | 30.3 | 22.2 | 15.8 | 14.4 |
| **1970, exercise** | Less exercise | 20.0 | 20.5 | 21.4 | 18.9 | 14.5 | 13.6 |
|  | no change | 44.2 | 53.9 | 43.7 | 54.4 | 69.7 | 72.0 |
|  | More | 35.8 | 25.7 | 34.9 | 26.8 | 15.8 | 14.4 |
| **1970, alcohol** | More alcohol | 28.6 | 20.3 | 26.3 | 22.2 | 14.5 | 13.6 |
|  | no change | 62.5 | 71.5 | 65.5 | 69.2 | 69.7 | 72.0 |
|  | Less | 8.9 | 8.2 | 8.2 | 8.6 | 15.8 | 14.4 |
| **1970, fruit & veg** | Less fruit & veg | 16.8 | 11.0 | 14.5 | 13.6 | 14.5 | 13.6 |
|  | no change | 64.1 | 75.6 | 69.7 | 72.0 | 69.7 | 72.0 |
|  | More | 19.0 | 13.4 | 15.8 | 14.4 | 15.8 | 14.4 |
| **1958, sleep** | Less sleep | 21.9 | 10.5 | 16.1 | 17.2 | 9.4 | 11.1 |
|  | no change | 56.0 | 66.6 | 60.4 | 62.5 | 75.8 | 77.1 |
|  | More | 22.1 | 23.0 | 23.5 | 20.3 | 14.8 | 11.8 |
| **1958, exercise** | Less exercise | 18.9 | 18.6 | 19.4 | 16.0 | 9.4 | 11.1 |
|  | no change | 54.1 | 58.2 | 50.4 | 63.4 | 75.8 | 77.1 |
|  | More | 27.0 | 23.2 | 30.3 | 20.6 | 14.8 | 11.8 |
| **1958, alcohol** | More alcohol | 16.8 | 14.0 | 16.8 | 15.1 | 9.4 | 11.1 |
|  | no change | 71.1 | 74.3 | 72.2 | 70.4 | 75.8 | 77.1 |
|  | Less | 12.1 | 11.8 | 11.0 | 14.5 | 14.8 | 11.8 |
| **1958, fruit & veg** | Less fruit & veg | 13.2 | 7.5 | 9.4 | 11.1 | 9.4 | 11.1 |
|  | no change | 73.6 | 79.5 | 75.8 | 77.1 | 75.8 | 77.1 |
|  | More | 13.2 | 13.0 | 14.8 | 11.8 | 14.8 | 11.8 |
|  |  | Females | Males | High education (degree/higher) | Low education (GCSE/none) | White | Non-white |
| **1946, sleep** | Less sleep | 14.7 | 11.3 | 8.9 | 14.4 | 6.0 | 6.0 |
|  | no change | 70.1 | 82.8 | 75.7 | 79.0 | 86.1 | 82.1 |
|  | More | 15.2 | 5.9 | 15.4 | 6.7 | 7.9 | 11.9 |
| **1946, exercise** | Less exercise | 21.4 | 14.9 | 17.4 | 15.3 | 6.0 | 6.0 |
|  | no change | 55.7 | 63.8 | 54.9 | 63.5 | 86.1 | 82.1 |
|  | More | 22.9 | 21.4 | 27.7 | 21.2 | 7.9 | 11.9 |
| **1946, alcohol** | More alcohol | 15.0 | 6.7 | 14.1 | 9.9 | 6.0 | 6.0 |
|  | no change | 75.0 | 84.8 | 72.2 | 82.2 | 86.1 | 82.1 |
|  | Less | 10.0 | 8.5 | 13.8 | 7.9 | 7.9 | 11.9 |
| **1946, fruit & veg** | Less fruit & veg | 8.4 | 3.6 | 6.0 | 6.0 | 6.0 | 6.0 |
|  | no change | 80.6 | 83.9 | 86.1 | 82.1 | 86.1 | 82.1 |
|  | More | 11.1 | 12.6 | 7.9 | 11.9 | 7.9 | 11.9 |

**Supplementary Table 2.** Behaviors pre and during COVID-19 lockdown: percentages in each cohort and risk factor group.

|  |  | <u>Cohort</u> |  |  |  |  | <u>Gender</u> |  | <u>Education</u> |  | <u>Ethnicity</u> |  |
| --- | --- | --- | --- | --- | --- | --- | --- | --- | --- | --- | --- | --- |
| **Outcome** | Whole sample | 2001 | 1990 | 1970 | 1958 | 1946 | Females | Males | High (degree /higher ) | Low (GCSE /none) | White | Non-white |
| **Sleep, pre** (hours/night) |  |  |  |  |  |  |  |  |  |  |  |  |
| **1** | 0.0% | 0.1% | 0.0% | 0.1% | 0.0% | 0.0% | 0.0% | 0.1% | 0.0% | 0.0% | 0.0% | 0.0% |
| **2** | 0.1% | 0.0% | 0.1% | 0.1% | 0.3% | 0.0% | 0.2% | 0.1% | 0.0% | 0.2% | 0.0% | 0.0% |
| **3** | 0.6% | 0.5% | 0.5% | 1.1% | 0.4% | 0.7% | 0.6% | 0.6% | 0.5% | 0.6% | 0.5% | 0.1% |
| **4** | 1.8% | 1.8% | 1.3% | 2.7% | 1.6% | 1.0% | 2.0% | 1.6% | 1.3% | 1.9% | 1.3% | 4.2% |
| **5** | 6.2% | 4.1% | 3.3% | 7.3% | 6.6% | 7.7% | 6.5% | 5.9% | 4.8% | 7.8% | 3.3% | 5.9% |
| **6** | 22.0% | 16.3% | 20.1% | 22.4% | 23.7% | 25.2% | 21.9% | 22.2% | 19.6% | 23.9% | 17.7% | 20.0% |
| **7** | 33.1% | 24.2% | 37.7% | 37.4% | 33.4% | 31.2% | 32.7% | 33.5% | 36.7% | 29.7% | 30.0% | 29.8% |
| **8** | 29.2% | 34.9% | 30.9% | 25.1% | 28.9% | 29.4% | 28.5% | 29.9% | 30.2% | 28.8% | 34.0% | 28.1% |
| **9** | 4.9% | 11.7% | 4.4% | 3.0% | 4.0% | 3.5% | 5.8% | 4.1% | 5.1% | 4.9% | 8.8% | 6.9% |
| **10** | 1.6% | 4.8% | 1.3% | 0.6% | 1.0% | 1.3% | 1.5% | 1.7% | 1.4% | 1.8% | 3.3% | 3.4% |
| **11** | 0.2% | 0.7% | 0.1% | 0.0% | 0.1% | 0.0% | 0.2% | 0.1% | 0.2% | 0.1% | 0.4% | 1.0% |
| **12** | 0.2% | 0.7% | 0.3% | 0.2% | 0.1% | 0.0% | 0.2% | 0.2% | 0.2% | 0.3% | 0.5% | 0.2% |
| **13** | 0.0% | 0.0% | 0.0% | 0.0% | 0.0% | 0.0% | 0.0% | 0.0% | 0.0% | 0.0% | 0.0% | 0.0% |
| **≥14** | 0.0% | 0.2% | 0.0% | 0.0% | 0.0% | 0.0% | 0.0% | 0.0% | 0.0% | 0.1% | 0.1% | 0.4% |
| **Sleep, during** (hours/night) |  |  |  |  |  |  |  |  |  |  |  |  |
| **1** | 0.1% | 0.3% | 0.0% | 0.1% | 0.2% | 0.0% | 0.1% | 0.1% | 0.1% | 0.3% | 0.2% | 0.0% |
| **2** | 0.4% | 0.6% | 0.3% | 0.7% | 0.1% | 0.1% | 0.5% | 0.3% | 0.2% | 0.1% | 0.5% | 0.1% |
| **3** | 0.8% | 0.8% | 0.2% | 1.2% | 0.6% | 0.9% | 0.8% | 0.8% | 0.6% | 1.1% | 0.6% | 0.3% |
| **4** | 3.4% | 3.1% | 2.4% | 3.9% | 3.6% | 3.1% | 3.8% | 2.9% | 2.3% | 4.3% | 2.5% | 5.5% |
| **5** | 7.9% | 4.8% | 6.3% | 9.3% | 7.9% | 9.6% | 8.8% | 6.9% | 6.3% | 9.7% | 5.0% | 8.3% |
| **6** | 18.1% | 10.8% | 16.2% | 19.2% | 19.9% | 21.1% | 18.5% | 17.7% | 16.2% | 20.1% | 12.2% | 19.2% |
| **7** | 25.6% | 12.1% | 23.5% | 27.9% | 28.0% | 32.0% | 24.5% | 26.8% | 26.2% | 24.0% | 17.8% | 11.3% |
| **8** | 27.3% | 24.8% | 32.3% | 26.2% | 28.2% | 26.4% | 26.1% | 28.6% | 29.8% | 25.5% | 28.1% | 27.3% |
| **9** | 9.9% | 20.6% | 11.6% | 8.2% | 8.4% | 4.4% | 10.2% | 9.7% | 11.1% | 8.8% | 17.7% | 10.2% |
| **10** | 5.1% | 15.6% | 5.8% | 2.7% | 2.9% | 2.3% | 5.1% | 5.0% | 5.6% | 4.7% | 11.5% | 10.9% |
| **11** | 0.6% | 3.0% | 0.3% | 0.2% | 0.1% | 0.0% | 0.6% | 0.5% | 0.6% | 0.6% | 1.8% | 1.7% |
| **12** | 0.7% | 3.0% | 0.8% | 0.2% | 0.1% | 0.1% | 0.9% | 0.5% | 0.9% | 0.6% | 1.8% | 4.0% |
| **13** | 0.0% | 0.2% | 0.0% | 0.0% | 0.0% | 0.0% | 0.0% | 0.0% | 0.0% | 0.1% | 0.1% | 0.4% |
| **≥14** | 0.1% | 0.4% | 0.2% | 0.1% | 0.0% | 0.0% | 0.1% | 0.1% | 0.1% | 0.1% | 0.3% | 0.7% |
| **Sleep, pre** (atypical, <6 or >9) | 10.7% | 12.9% | 6.9% | 12.0% | 10.0% | 10.8% | 11.2% | 10.2% | 8.5% | 12.7% | 9.5% | 15.2% |
| **Sleep, during** (atypical, <6 or >9) | 19.0% | 31.7% | 16.5% | 18.4% | 15.6% | 16.1% | 20.8% | 17.2% | 16.7% | 21.5% | 24.3% | 31.9% |

| **Outcome** | **Whole sample** | **2001** | **1990** | **1970** | **1958** | **1946** | **Females** | **Males** | **High (degree** | **Low (GCSE /none)** | **White** | **Non-white** |
| --- | --- | --- | --- | --- | --- | --- | --- | --- | --- | --- | --- | --- |
| **Exercise, pre (times/week)** |  |  |  |  |  |  |  |  |  |  |  |  |
| 0 | 19.6% | 16.6% | 20.2% | 19.5% | 19.2% | 22.7% | 22.0% | 17.0% | 15.6% | 23.6% | 18.0% | 18.5% |
| 1 | 9.4% | 12.2% | 11.8% | 10.1% | 7.7% | 6.8% | 10.3% | 8.4% | 10.1% | 8.8% | 12.4% | 9.2% |
| 2 | 13.8% | 15.9% | 15.3% | 13.4% | 13.2% | 12.1% | 14.5% | 13.0% | 15.7% | 12.9% | 15.2% | 19.4% |
| 3 | 15.7% | 16.3% | 17.8% | 16.6% | 14.5% | 14.5% | 17.1% | 14.2% | 16.6% | 14.0% | 17.3% | 14.6% |
| 4 | 11.8% | 13.0% | 10.7% | 11.0% | 11.2% | 13.8% | 11.3% | 12.4% | 11.9% | 11.4% | 11.6% | 15.6% |
| 5 | 13.6% | 12.3% | 14.8% | 15.2% | 14.0% | 10.6% | 12.1% | 15.0% | 14.1% | 13.3% | 13.8% | 11.1% |
| 6 | 4.7% | 5.7% | 4.1% | 3.9% | 5.3% | 4.5% | 3.7% | 5.8% | 4.8% | 4.4% | 5.1% | 5.3% |
| 7 | 11.5% | 7.8% | 5.3% | 10.2% | 14.9% | 15.1% | 9.0% | 14.1% | 11.2% | 11.6% | 6.7% | 6.4% |
| **Exercise, during (times/week)** |  |  |  |  |  |  |  |  |  |  |  |  |
| 0 | 20.7% | 16.5% | 20.1% | 19.9% | 21.6% | 24.7% | 21.6% | 19.8% | 16.0% | 25.1% | 17.4% | 21.8% |
| 1 | 9.3% | 13.9% | 12.4% | 9.3% | 7.4% | 6.0% | 9.4% | 9.1% | 10.1% | 8.7% | 12.7% | 17.3% |
| 2 | 10.9% | 13.0% | 10.8% | 10.7% | 9.8% | 11.3% | 11.7% | 10.1% | 10.7% | 11.2% | 12.4% | 10.1% |
| 3 | 12.9% | 13.8% | 18.9% | 13.9% | 10.5% | 10.6% | 13.5% | 12.2% | 13.7% | 12.2% | 16.1% | 14.6% |
| 4 | 10.7% | 12.6% | 11.5% | 10.1% | 9.9% | 11.0% | 10.1% | 11.4% | 11.4% | 9.7% | 11.4% | 18.0% |
| 5 | 12.8% | 12.1% | 10.9% | 15.2% | 13.4% | 10.0% | 13.3% | 12.4% | 13.9% | 11.6% | 12.1% | 8.7% |
| 6 | 7.0% | 7.6% | 5.6% | 6.4% | 8.0% | 6.9% | 6.9% | 7.2% | 7.6% | 6.1% | 7.0% | 4.7% |
| 7 | 15.6% | 10.4% | 9.8% | 14.5% | 19.3% | 19.5% | 13.4% | 17.8% | 16.7% | 15.2% | 10.9% | 4.9% |
| **Exercise, pre (<3 times/week)** | 28.9% | 28.8% | 32.0% | 29.7% | 26.9% | 29.5% | 32.4% | 25.4% | 25.7% | 32.3% | 30.4% | 27.7% |
| **Exercise, during (<3 times/week)** | 30.0% | 30.4% | 32.5% | 29.2% | 29.1% | 30.8% | 31.1% | 28.9% | 26.1% | 33.9% | 30.1% | 39.1% |
| **Alcohol, pre (times/week)** |  |  |  |  |  |  |  |  |  |  |  |  |
| 4 or more times a week | 18.0% | 8.4% | 7.3% | 15.2% | 23.7% | 28.5% | 13.8% | 22.3% | 19.7% | 16.7% | 8.7% | 3.0% |
| 2-3 times a week | 26.5% | 26.2% | 19.6% | 29.1% | 28.2% | 24.2% | 24.9% | 28.0% | 30.9% | 24.0% | 25.0% | 13.3% |
| 2-4 times per month | 21.4% | 29.7% | 27.6% | 22.2% | 17.5% | 15.2% | 22.2% | 20.6% | 21.6% | 20.4% | 30.3% | 19.0% |
| Monthly or less | 17.2% | 20.4% | 25.0% | 17.4% | 14.4% | 13.6% | 21.0% | 13.3% | 14.6% | 19.5% | 23.0% | 16.5% |
| Never | 16.9% | 15.3% | 20.5% | 16.1% | 16.2% | 18.5% | 18.0% | 15.8% | 13.3% | 19.4% | 13.0% | 48.3% |
| **Alcohol, during (times/week)** |  |  |  |  |  |  |  |  |  |  |  |  |
| 4 or more times a week | 23.8% | 8.9% | 14.6% | 25.5% | 29.2% | 32.0% | 20.3% | 27.4% | 26.2% | 21.8% | 11.8% | 8.2% |
| 2-3 times a week | 24.2% | 19.5% | 23.9% | 27.0% | 25.3% | 22.5% | 23.4% | 25.0% | 26.8% | 23.1% | 23.2% | 9.4% |
| 2-4 times per month | 17.0% | 24.0% | 20.4% | 16.7% | 13.8% | 14.2% | 17.3% | 16.6% | 16.5% | 16.2% | 24.2% | 9.3% |
| Monthly or less | 13.0% | 19.9% | 15.9% | 11.9% | 10.7% | 10.6% | 15.3% | 10.7% | 12.3% | 14.0% | 18.7% | 15.3% |
| Never | 22.0% | 27.7% | 25.2% | 19.0% | 20.9% | 20.7% | 23.7% | 20.2% | 18.2% | 24.8% | 22.1% | 57.8% |
| **Alcohol, pre (high risk drinking)** | 19.2% | 32.6% | 16.0% | 17.3% | 17.2% | 14.2% | 14.0% | 24.3% | 21.0% | 18.6% | 27.4% | 13.6% |
| **Alcohol, during (high risk drinking)** | 16.8% | 13.0% | 12.7% | 21.7% | 17.4% | 14.6% | 11.8% | 21.9% | 18.1% | 16.7% | 13.3% | 10.1% |

| Outcome | Whole sample | 2001 | 1990 | 1970 | 1958 | 1946 | Females | Males | High (degree) | Low (GCSE /none) | White | Non-white |
| --- | --- | --- | --- | --- | --- | --- | --- | --- | --- | --- | --- | --- |
| **Fruit & veg, pre (portions/day)** |  |  |  |  |  |  |  |  |  |  |  |  |
| 0 | 1.6% | 3.5% | 1.5% | 1.9% | 1.1% | 0.3% | 1.2% | 2.0% | 1.0% | 1.8% | 2.4% | 4.3% |
| 1 | 8.5% | 14.0% | 8.6% | 10.0% | 7.4% | 3.1% | 5.6% | 11.4% | 7.3% | 8.8% | 11.4% | 13.2% |
| 2 | 16.3% | 21.9% | 16.9% | 15.5% | 15.9% | 12.9% | 14.7% | 18.0% | 15.4% | 17.0% | 19.7% | 21.0% |
| 3 | 23.3% | 25.6% | 28.4% | 22.5% | 22.2% | 21.2% | 23.5% | 23.2% | 22.8% | 24.4% | 26.7% | 28.5% |
| 4 | 18.8% | 16.9% | 16.7% | 17.2% | 17.8% | 26.4% | 19.4% | 18.2% | 19.0% | 19.3% | 17.5% | 10.8% |
| 5 | 19.4% | 10.8% | 20.1% | 21.7% | 21.2% | 19.6% | 21.1% | 17.7% | 21.5% | 17.7% | 15.1% | 13.4% |
| 6 | 12.1% | 7.2% | 7.8% | 11.2% | 14.5% | 16.5% | 14.6% | 9.6% | 12.9% | 11.0% | 7.2% | 8.8% |
| **Fruit & veg, during (portions/day)** |  |  |  |  |  |  |  |  |  |  |  |  |
| 0 | 1.7% | 3.5% | 2.5% | 2.1% | 0.9% | 0.2% | 1.2% | 2.2% | 1.4% | 2.0% | 3.0% | 3.3% |
| 1 | 8.7% | 12.1% | 9.4% | 11.5% | 7.5% | 3.0% | 6.3% | 11.2% | 7.2% | 9.1% | 10.2% | 16.3% |
| 2 | 15.0% | 18.7% | 14.9% | 13.8% | 16.1% | 11.4% | 13.8% | 16.2% | 13.9% | 16.1% | 17.0% | 17.9% |
| 3 | 22.6% | 22.0% | 25.0% | 23.0% | 21.6% | 22.9% | 23.1% | 22.1% | 21.1% | 24.6% | 23.0% | 25.3% |
| 4 | 19.0% | 19.4% | 19.0% | 18.1% | 16.8% | 24.7% | 19.1% | 19.0% | 19.9% | 18.7% | 19.7% | 15.7% |
| 5 | 19.2% | 14.2% | 18.7% | 19.6% | 21.2% | 19.8% | 19.9% | 18.5% | 21.8% | 17.6% | 16.7% | 12.2% |
| 6 | 13.7% | 10.0% | 10.4% | 12.0% | 15.9% | 18.0% | 16.6% | 10.8% | 14.8% | 12.1% | 10.3% | 9.2% |
| ***Fruit & veg, pre (< 3 portions a day)*** | 26.4% | 39.4% | 27.0% | 27.4% | 24.3% | 16.3% | 21.4% | 31.3% | 23.8% | 27.6% | 33.5% | 38.5% |
| ***Fruit & veg, during (< 3 portions a day)*** | 25.4% | 34.4% | 26.9% | 27.3% | 24.5% | 14.7% | 21.3% | 29.6% | 22.5% | 27.1% | 30.3% | 37.5% |

Notes:
High risk drinking is consuming more than 14 drinks a week or more than 5 drinks in a typical drinking day.
Measures in italics are those used in the main analyses shown in Figure 2.

